# Endovascular Treatment Combined with Remote Ischemic Postconditioning in Patients with Acute Ischemic Stroke Improves the Prognosis, a Multicenter, Randomized, Prospective Trial(EnTRIPS): study protocol for a randomized clinical trial

**DOI:** 10.1101/2025.03.20.25324340

**Authors:** Wanying Chen, Meng wei, Yawen Cheng, Fude Liu, Xiangyu Lei, Mengmeng Li, Panpan Zhang, Guoliang Li, Guogang Luo

**Author notes:** Corresponding author: Guoliang Li, Guogang Luo.

## Abstract

Endovascular therapy is a crucial treatment for patients with acute ischemic stroke(AIS), however many individuals experiencing cerebral ischemia-reperfusion (I/R) injury experience poor prognosis. Improving the clinical effectiveness of endovascular therapy remains a major clinical challenge, particularly in enhancing neuroprotection. Remote ischemic postconditioning (RIPC) has demonstrated potential in improving outcomes for AIS patients, especially those undergoing intravenous thrombolysis. However, the specific neuroprotective effects of RIPC in patients who have successfully undergone endovascular recanalization remain unexplored. This study aims to assess the safety and efficacy of RIPC in patients receiving endovascular therapy with successful recanalization.

In this multicenter study, AIS patients from several tertiary hospitals who underwent endovascular therapy with successful recanalization were randomly divided into the RIPC group or control group. The RIPC group received postoperative distal ischemia adaptation twice daily until discharge for at least 7 days, while the control received standard care without RIPC intervention. The primary outcome measure was the proportion of patients achieving a favorable outcome (mRS ≤ 2) at 3 months post-surgery. We hypothesize that integrating RIPC with endovascular therapy will significantly enhance the functional prognosis of AIS patients, potentially leading to better functional prognosis.

## 1. Introduce

Ischemic stroke is characterized by a high incidence, recurrence and disability rate, making it the second leading cause of death in the world, bringing huge living and economic burden to patients and society^[1,2]^. Currently, intravenous thrombolytic (IVT) within 4.5h is the preferred treatment method for acute ischemic stroke (AIS). Although IVT therapy has been widely adopted and the technology is well-established, the number of patients who arrive at the hospital within this time frame and are suitable for IVT is very limited ^[3]^. In addition, the proportion of recanalization through IVT is generally low in patients with large vascular occlusive stroke, so endovascular therapy (ET) is required to increase the rate of vascularization and reduce mortality and disability. However, although approximately 80% of blocked vessels can be recanalized after ET, only about 50% of patients achieve functional independence 90 days after treatment, and more than 15% of patients even die ^[4,5]^. There are many reasons for poor prognosis, among which ischemia/reperfusion injury is the most important. Studies have shown ^[6,7]^ that although restoration of blood flow is the main goal of treatment for AIS, if it occurs too late, it can lead to more severe damage than if there is no revascularization. Therefore, the development of new therapies to improve the prognosis of AIS patients remain an urgent task for many physicians.

Ischemic conditioning (IC) is when an organ or tissue is subjected to repeated, temporary, and non-fatal Ischemic stimulation that causes tolerance to the body or distant organs and reduces subsequent Ischemic injury. In recent years, there have been more and more studies on the relationship between remote ischemic postconditioning (RIPC) and stroke. RIPC can not only improve the neurological injury of AIS patient^[8,9]^, but also reduce the risk of stroke recurrence and death of non-AIS^[10]^, which is an effective method for secondary prevention. RIPC is also safe and effective as an adjunct to IVT^[11]^. However, there are few studies on the efficacy and safety of RIPC combined with ET for AIS, so the purpose of this study is to explore the efficacy and safety of RIPC combined with ET for AIS.

## Materials and Methods

### Study design

The EnTRIPS study (Endovascular Treatment Combined with Remote Ischemic Postconditioning in Patients with Acute Ischemic Stroke Improves the Prognosis) is a multicenter, randomized, prospective Trial(registered on clinicaltrials. gov with NCT04581759) performed at the following tertiary hospitals in China: the First Affiliated Hospital of Xi’an Jiaotong University, the Second Affiliated Hospital of Xi’an Jiao Tong University, Provincial People’s Hospital of Shaanxi, Xi’an No. 3 Hospital, Xi’an Gao xin Hospital, Xi dian group Hospital, Xi ‘an Ninth Hospital and Xian Yang Hospital of Yan ‘an University. The protocol was approved by the Ethics Committee of the First Affiliated Hospital of Xi’an Jiao Tong University. All participants or their legally authorized representatives provided written informed consent. Figure 1 shows the schedule of enrollment, intervention, and assessments.

**Figure 1.**
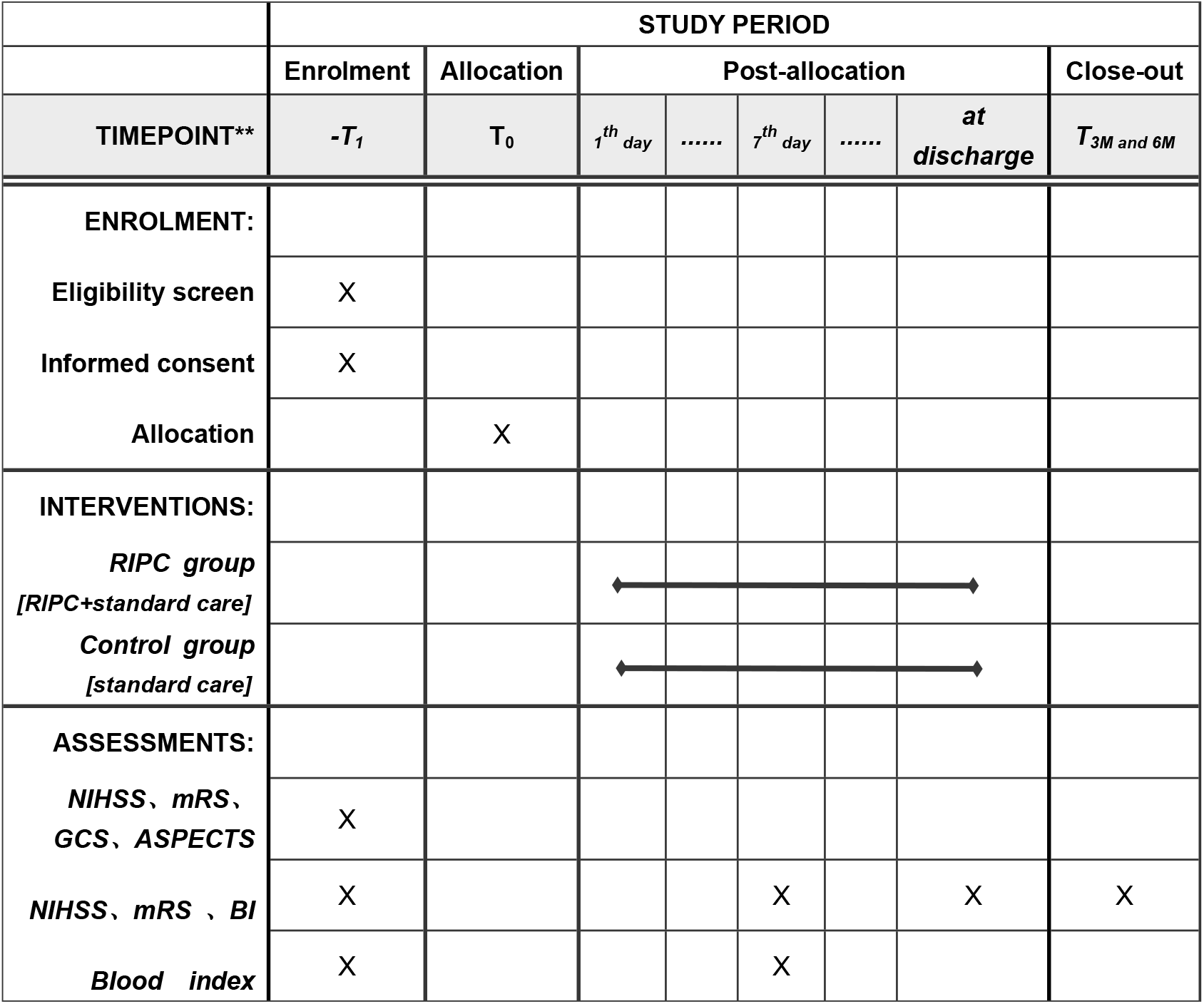
Schedule of enrollment, intervention, and assessments. NIHSS National Institute of Health Stroke Scale, mRS modified Rankin Scale, ASPECTS Alberta Stroke Program Early CT score, GCS Glasgow,BI Barthel Index.

### Participant selection

The subjects for this study will be recruited from 8 hospitals in Xi ‘an, and all patients will be evaluated by neurologist before and after surgery to determine whether patients conform to the inclusion and exclusion criteria before offering them the opportunity to participate in the study.

#### Inclusion criteria

The study inclusion criteria were as follows:

1. AIS patients with large vessel occlusion (internal carotid artery system and vertebrobasilar artery system) within 24 hours of onset, who underwent endovascular treatment (mechanical thrombectomy, arterial thrombolysis, balloon dilatation, stent implantation) and successful recanalization were defined as: The modified Thrombolysis in Cerebral infarction (mTICI) criteria were ≥ 2B, and the criteria for endovascular intervention were in line with the indications and condictions set out in the Chinese Guidelines for Early Endovascular Intervention in Acute Ischemic Stroke 2018.
2. Pre-onset modified Rankin Scale (mRS) score ≤1;
3. Alberta Stroke Program Early CT score (ASPECTS)≥6 on admission;
4. NIHSS score ≥6 on admission;
5. Provision of written informed consent.

#### Exclusion criteria

Participants who met any of the following criteria were excluded from the study:

1. CT or MRI scan showed significant midline deviation and the mass effect;
2. Glasgow (GCS) score ≤8;
3. Failure to complete 3-months and 6-months follow-up;
4. Severe cardiac, liver, or kidney disease, malignancy, severe coagulation dysfunction, severe anemia and systemic organ dysfunction;
5. pregnant or breastfeeding women, or patients with moderate-severe mental disorders or dementia;
6. Severe soft tissue injuries, fractures, thrombosis and other known peripheral vascular lesions of the upper limbs, active visceral hemorrhage, acute stage of fundus hemorrhage, cerebral aneurysm or cerebral arteriovenous malformation, and other unsuitable for bilateral upper arm compression.

### Randomization

All patients were randomized by the Clinical Research Center of the First Affiliated Hospital of Xi’an Jiaotong University using SAS software, and the inclusion category of each patient was determined to receive RIPC combined with standard stroke unit therapy or stroke unit therapy alone. The specific methods were as follows: 10 patients in each block were allocated to the RIPC group or control group in a 1:1 ratio. In a center, when one enrolled case is obtained, a block of 10 consecutive case grouping schemes can be obtained. When the center completes the records of these 10 patients, the next block will be contested. The enrolled physicians interviewed the participants or their families and signed informed consent. The on-call nurses were not blinded to the treatment instructions, but they were not involved in data analysis or follow-up clinical scores. The observers assessing the clinical outcome were blinded to the treatment assignment.

### Procedures

We recruited AIS patients who underwent ET within 24 hours of symptom onset and had successful recanalization, with recanalization defined as modified Thrombolysis in Cerebral Ischemia (mTICI) ≥ 2B. Baseline demographic, clinical, and laboratory information was collected during surgery. Eligible participants were randomly divided into the RIPC group or control group and provided signed informed consent.

Patients in the RIPC group underwent 5 cycles of RIPC consisting of automatic alternating cuff inflation (to 180 mm Hg for 5 minutes) and deflation (for 3 minutes) on both arms by a user-friendly device (IPC-906X; Beijing Renqiao Institute of Neuroscience, Beijing, China). The total procedure time was 40 minutes, twice per day, for at least 7 days during hospitalization, and may be interrupted due to temporary intolerance, bleeding spots on the upper arm, etc. The first RIPC treatment was started within 6 hours after surgery. When participants were not suitable for bilateral treatment (for example, because they were receiving intravenous fluids, using blood pressure monitors, etc.), it was performed in the unilateral upper extremity.

The process was completed with the help of a nurse at the hospital, and blood pressure and heart rate were recorded before and after each treatment. Participants can stop the process at any time if they feel uncomfortable. The control group underwent no inflation or deflation. All patients received active treatment and best practice management in accordance with guidelines for the treatment of acute stroke.

### Blood sample

Venous blood was collected at three time points: before surgery and the seventh day after surgery. Blood sample is processed at 3000r, 4°C for 10min, and then stored as serum/plasma at −80°C.

### Outcome measure

#### Primary outcome measure

The primary outcome measure will be the proportion of patients who are functionally independent (defined as mRS Score ≤2) 3M after surgery, which will be compared between the intervention and control groups.

#### Secondary outcome measures

(1) The proportion of symptomatic cerebral hemorrhage within 72h after surgery; (2) Neurological function scores of patients at discharge and 7 days, 3M and 6M after surgery; (3) The incidence and mortality of stroke recurrence and other related cerebrovascular diseases at discharge and 7 days, 3M and 6M after surgery.

#### Other outcome measures

The differences of key laboratory examination indicators between the two groups.

### Sample size estimates

Based on previous studies^[12-14]^, the short-term good prognosis rate of AIS patients with ET and successful recanalization was about 48%. And according to our previous study^[11]^, the ratio of the end point event rate of the RIPC group to the end point event rate of the conventional group was 1.38 (meaning that combined treatment with RIPC is expected to further increase the effective rate of 18% on the basis of conventional treatment). In addition, considering a significance level of 5% and power of 80%, the final sample size was calculated to be 230 (115 per group). With a proposed 10% dropout rate, 254(127 per group) participants will be recruited for this study.

### Statistical analysis

Continuous variables will be express as the median (lower quartile-high quartile) and categorical variables will be express as frequencies (percentages). For group comparisons of data with a normal distribution, we utilize t-test. For group comparisons of skewed variables, then a logarithmic transformation or non-parametric alternatives will be used to analyze the data.

To analyze the neurological function (mRS, NIHSS, and BI) score, we will evaluate the odds ratio for a shift in the direction of a better outcome on the neurological function prognosis in both groups. The ratio will be estimated using logistic regression and all possible cutoff values on mRS, NIHSS and BI will be calculated. Analyses will be done on the full data, and a sub-analysis will be performed excluding the influence of age, etiology and other factors on the treatment effect of RIPC.

### Current status

Our study enrolled the first patient on April 13, 2021, and is expected to complete enrollment by the end of May 2025, with 3-month follow-up and data collation by the end of August 2025.

## Discussion

To the best of our knowledge, this is a pioneering clinical trial to investigate the role of RIPC in AIS patients receiving ET with successful recanalization. Previous studies have shown good results for RIPC in AIS patients alone and those who have undergone IVT. Another trial (registration number NCT04977869 ^[15]^) similar to our study is currently being conducted to investigate the effect of RIPC in AIS patients receiving ET with successful recanalization, however our study was the first to register and start enrolling patients and and there are differences as follow: 1.We only recruited patients with ET and successful recanalization, and they also enrolled patients with ET and failed to recanalization; 2.We recruited AIS patients with large vessel occlusion in anterior and posterior circulation, and they only enrolled AIS patients with large vessel occlusion in anterior circulation; 3. Our RIPC treatment was started within 6 hours after surgery with an inflation pressure of both upper arms, and their RIPC treatment was started after ET with an inflation pressure of one upper arms.

There are no clear conclusions about the effects of this intervention in this group.The main concern of this patient group during use was whether the RIPC treatment would increase blood pressure and cerebral perfusion and affect cerebral hydrodynamics, leading to cerebral hyperperfusion. Zhao et al.^[16]^ measured blood pressure, intracranial pressure, cerebral perfusion pressure, bilateral middle cerebral artery systolic blood flow velocity peak, pulse index and other indicators before, during and after RIPC treatment for self-comparison, proving that RIPC is safe and feasible for AIS patients receiving ET. In clinical practice, it is not clear what mode of RIPC treatment is needed to make the patient’s prognosis better. In this study, a relatively common method was adopted, namely, the band was inflated to 180 mmHg, the upper arms were pressurized for 5 min, and the treatment was relaxed for 3 min, and the treatment lasted 5 cycles at a time. Choosing the appropriate time window for RIPC treatment is another key issue. In the process of brain I/R, many reversible injuries are transformed into irreversible injuries during the early stage of brain I/R, so early RIPC treatment after recanalization may be more effective in playing a neuroprotective role. Ren et al. ^[17]^ proved with a rat stroke model that rapid postconditioning had better neuroprotective effects than delayed postconditioning in some RIPC modes.Therefore, in order to maximize the protective effect of RIPC, this study applied double upper arm compression therapy as soon as possible within 6 h after recanalization.

The main outcome of the study was the ratio of 90 d functional independence (mRS≤2) between the RIPC group and the control group. It will be possible to recommend the implementation of RIPC therapy in AIS patients with emergency cerebrovascular recanalization, and provide a basis for studies on the application and mechanism of RIPC therapy in such patients.

## Data Availability

No datasets were generated or analysed during the current study.

## Acknowledgements

We sincerely thank the participants in the clinical trial and the doctors and nurses in the neurology department for their help and support.

## Funding

This study was supported by the Clinical Research Award of the First Affiliated Hospital of Xi’an Jiaotong University, China(No. XJTU1AF2021CRF-003); the Clinical Research Award of the First Affiliated Hospital of Xi’an Jiaotong University, China(No. XJTU1AF-CRF-2020-015).

## Declarations

### Ethics approval and consent to participate

This study has been reviewed and approved by the Medical Ethics Committee of the First Affiliated Hospital of Xi ‘an Jiaotong University(No.XJTU1AF2021LSK–093). All participants will be informed of the purpose of the study and sign an informed consent form.

## Conflicts of interest

No potential conflicts of interest related to this article were reported.

## Notes

### Competing Interest Statement

The authors have declared no competing interest.

### Clinical Trial

NCT04581759

### Funding Statement

Yes

### Author Declarations

The protocol was approved by the Ethics Committee of the First Affiliated Hospital of Xi’an Jiao Tong University(NO:XJTU1AF2021LSK-093). All participants or their legally authorized representatives provided written informed consent.

